# Safety and Immunogenicity of CpG 1018 and Aluminium Hydroxide-Adjuvanted SARS-CoV-2 S-2P Protein Vaccine MVC-COV1901: A Large-Scale Double-Blind, Randomised, Placebo-Controlled Phase 2 Trial

**DOI:** 10.1101/2021.08.05.21261532

**Authors:** Hsieh Szu-Min, Ming-Che Liu, Yen-Hsu Chen, Wen-Sen Lee, Shinn-Jang Hwang, Shu-Hsing Cheng, Wen-Chien Ko, Kao-Pin Hwang, Ning-Chi Wang, Yu-Lin Lee, Yi-Ling Lin, Shin-Ru Shih, Chung-Guei Huang, Chun-Che Liao, Jian-Jong Liang, Chih-Shin Chang, Charles Chen, Chia En Lien, I-Chen Tai, Tzou-Yien Lin

## Abstract

**Background:** We have assessed the safety and immunogenicity of the COVID-19 vaccine MVC-COV1901, a recombinant protein vaccine containing prefusion-stabilized spike protein S-2P adjuvanted with CpG 1018 and aluminium hydroxide.

**Methods:** This is a phase 2, prospective, randomised, double-blind, placebo-controlled, and multi-centre study to evaluate the safety, tolerability, and immunogenicity of the SARS-CoV-2 vaccine candidate MVC-COV1901. The study comprised 3,844 participants of ≥ 20 years who were generally healthy or with stable pre-existing medical conditions. The study participants were randomly assigned in a 6:1 ratio to receive either MVC-COV1901 containing 15 μg of S-2P protein or placebo containing saline. Participants received two doses of MVC-COV1901 or placebo, administered 28 days apart via intramuscular injection. The primary outcomes were to evaluate the safety, tolerability, and immunogenicity of MVC-COV1901 from Day 1 (the day of first vaccination) to Day 57 (28 days after the second dose). Immunogenicity of MVC-COV1901 was assessed through geometric mean titres (GMT) and seroconversion rates (SCR) of neutralising antibody and antigen-specific immunoglobulin. This clinical trial is registered at ClinicalTrials.gov: NCT04695652.

**Findings:** From the start of this phase 2 trial to the time of interim analysis, no vaccine-related Serious Adverse Events (SAEs) were recorded. The most common solicited adverse events across all study participants were pain at the injection site (64%), and malaise/fatigue (35%). Fever was rarely reported (<1%). For all participants in the MVC-COV1901 group, at 28 days after the second dose against wild type SARS-CoV-2 virus, the GMT was 662·3 (408 IU/mL), the GMT ratio was 163·2, and the seroconversion rate was 99·8%.

**Interpretation:** MVC-COV1901 shows good safety profiles and promising immunogenicity responses. The current data supports MVC-COV1901 to enter phase 3 efficacy trials and could enable regulatory considerations for Emergency Use Authorisation (EUA).

**Funding:** Medigen Vaccine Biologics Corporation and Taiwan Centres for Disease Control.

## Introduction

In December 2019, the Coronavirus Disease (COVID-19) was first reported in Wuhan, China. Identified as a SARS related coronavirus, it was named SARS-CoV-2. The disease spread with such ferocity, the WHO declared a global pandemic on 11 March 2020. Despite the unprecedented pace of vaccine development, COVID-19 continues to wreak havoc, with over 170 million infections and 3·7 million deaths worldwide as of June 2021.^3^ Even though several vaccines that have obtained Emergency Use Authorisation (EUA), using mRNA platform or adenovirus vector-based technology, the limitations and global supply of available vaccines are not enough to end this pandemic. The addition of traditional recombinant subunit protein vaccines could bolster current vaccination efforts.

MVC-COV1901 is a recombinant protein subunit vaccine based on the stabilised prefusion SARS-CoV-2 spike protein S-2P adjuvanted CpG 1018 and aluminium hydroxide.^4,5^ The S-2P protein was developed by McLellan and Graham at the National Institute of Allergy and Infectious Diseases (NIAID), U.S.A, and the synthetic oligodeoxynucleotide CpG 1018 adjuvant is developed by Dynavax.^4^ Using animal models, we have demonstrated a total protection in hamsters challenged with wild type SARS-COV-2.^6^ The result of our phase 1 trial confirmed that MVC-COV1901 is well tolerated and elicits robust immune responses in both T-cell & B-cell immunity.^7^ For this phase 2 trial, we proceeded with the 15 μg spike S-2P protein MVC-COV1901.

The purpose of this study is to assess the safety, tolerability, and immunogenicity of the MVC-COV1901 vaccine against placebo for adult participants who are healthy or with stable pre-existing medical conditions.

## Methods

### Trial design and oversight

This is an ongoing phase 2, prospective, randomised, double-blind, placebo-controlled (investigator/site staff and participants), multi-centre study. The study sites consisted of ten medical centres and one regional hospital in Taiwan. We planned to enrol approximately 3,700 adult participants. Participants were to be stratified into two age groups: younger adults (aged 20-64 years) and older adults (aged 65 years and over). The older adults were to make up 20% of the participants.

The trial protocol and informed consent form were approved by Taiwan Food and Drug Administration (TFDA) and the ethics committees at the conducting sites (Main review Institutional. Review Board [IRB]: Chang Gung Medical Foundation; sub Review IRBs: National Taiwan University Hospital; Taipei Veterans General Hospital, Tri-Service General Hospital, Taipei Medical University Hospital, Taipei Municipal Wanfang Hospital, Taoyuan General Hospital MOHW, China Medical University Hospital, Changhua Christian Hospital, National Cheng Kung University Hospital, and Kaoshiung Medical University Hospital). An Independent Data Monitoring Committee (IDMC) was established to monitor data safety and trial conduct. The study complies with the protocol and statistical analysis plan. The protocol is available in the appendix. The trial was conducted in accordance with the principles of the Declaration of Helsinki and Good Clinical Practice (GCP) guidelines, and is registered at the ClinicalTrials.gov: NCT 04695652.

### Participants

Participants were male or female adults aged 20 years or over who were healthy or with stable pre-existing medical conditions. Female participants of child-bearing potential had to agree to take effective contraception. Those who travelled overseas within 14 days of screening, or those who planned to travel overseas in six months were excluded from screening. Participants must not have pre-specified medical conditions, including immunosuppressive illness, history of autoimmune disease, malignancy with risk to recur, bleeding disorder, uncontrolled human immunodeficiency virus (HIV), uncontrolled hepatitis B and C infections, SARS-CoV-1 or 2 virus infections, allergy to any vaccine, or serious medical conditions that would interfere with the study. A full list of inclusion and exclusion criteria is available in the protocol. All participants provided signed informed consent.

### Randomisation and masking

Participants were centrally randomised using an interactive web response system (IWRS). Participants received either MVC-COV1901 or placebo in a 6:1 ratio. The Safety Set of this interim analysis included all study participants who has received at least one dose of injection. The Per-Protocol Immunogenicity (PPI) Subset planned to include the first 1,090 randomised participants: 820 participants from the younger adult group and 270 participants from the older adult group.

This was a double-blind study in which participants and investigators were blinded to the study intervention. Since MVC-COV1901 and placebo are visually distinct, each trial site was assigned an independent study nurse, who was not involved in the preparation, dispensing, administration, and accountability of the study intervention. To ensure treatment blindness to all study staff and participants, study-specific training was conducted at every trial site.

### Procedures

Each participant received two doses of either MVC-COV1901 or placebo on Visit 2 (Day 1) and Visit 4 (Day 29) via intramuscular (IM) injection in the deltoid. The prefilled MVC-COV1901 contained 15 μg of CHO cell-derived S-2P protein adjuvanted with CpG 1018 750 μg and aluminium hydroxide 375 μg, as previously reported.^7^ The vaccine was produced at Medigen Vaccine Biologics Corp. facility and complied with the current good manufacturing practices (cGMP). The placebo was 0.5 mL of saline. The duration of the study was 209 days (29 days treatment period, 180 days follow-up). Participant data was collected via six on-site visits and three phone calls.

### Outcomes

#### Safety assessment

The primary safety endpoints were to evaluate the safety and tolerability of all participants receiving study intervention from Day 1 to Day 57 (28 days after the second dose). Vital signs were assessed before and after each injection. Participants were observed for at least 30 minutes after each injection to identify any immediate adverse events (AEs). After each injection, participants had to record solicited local and systemic AEs in their diary cards for up to seven days. Unsolicited AEs were recorded for 28 days following each injection. All other AEs, SAEs, adverse events of special interests (AESIs), and vaccine-associated enhanced disease (VAED) were recorded throughout the study period.

#### Immunogenicity assessment

The primary immunogenicity endpoint was to evaluate neutralising antibody titres on Day 57 in terms of geometric mean titres (GMT), GMT ratio, and seroconversion rate (SCR) in the PPI Subset. The secondary endpoint was to evaluate the anti-Spike IgG antibody titres on Day 29 (the day of second dose), Day 43 (14 days after the second dose), and Day 57 in the PPI Subset.

#### Laboratory methods

Wild type SARS-CoV-2 TCDC#4 strain (hCoV-19/Taiwan/4/2020, GISAID accession ID: EPI_ISL_411927) was titrated to calculate the TCID_50_. Vero E6 cells (1·2 × 10^5^ cells/well) were seeded in 96-well plates and incubated. The sera underwent a total of ten two-fold dilutions, starting from a 1:8 dilution to a final dilution of 1:8,192. Diluted sera were mixed with equal volume of 100 TCID_50_/50 μL of virus. The serum-virus mixture was incubated at 37 °C for one hour, and then added to the cell plates containing the Vero E6 cells, followed by a further incubation at 37 °C, 5% CO_2_ incubator for four to five days. The neutralising titre (NT_50_) was defined as the reciprocal of the highest dilution capable of inhibiting 50% of the cytopathic effect (CPE). The readouts came from quadruplicated tests that were calculated using the formula of the Reed-Muench method.

To facilitate conversion of GMT to WHO’s International Unit (IU/mL) or Binding Antibody Unit (BAU/mL), we purchased WHO international standard reference panels (NIBSC 20/136, NIBSC 20/268) from National Institute for Biological Standards and Control (NIBSC; Potters Bar, UK) for comparison.^8,9^ (Supplementary Table S1) Our methodology to convert GMT to IU/mL is as follows: first, a neutralising assay was developed to obtain the GMTs of the NIBSC sera. The GMTs were verified by three repeated tests. Using the correlation between the GMTs and its assigned IU/mL of each standardised NIBSC sera, we established an equation for converting GMT to IU/mL. We applied this equation to convert the GMTs of our study groups to IU/mL.

For the conversion of GMT to BAU/mL, the total serum anti-Spike IgG titres were measured using enzyme-linked immunosorbent assay (ELISA) customized 96-well plates coated with S-2P antigen, as previously reported.^7^ The WHO NIBSC 20/136 was tested with the same ELISA assay that was used in our study samples. The geometric mean of the anti-Spike IgG titre for NIBSC 20/163 was 10,960·9, which was calculated from seven repeated tests. Since NIBSC 20/136 was assigned as 1,000 BAU/mL, a conversion factor of 0·0912 (1,000/10,960·9) was established to predict BAU/mL from anti-Spike IgG titres. The raw data for the conversion of anti-Spike IgG GMT to BAU/mL is provided in the Supplementary Table S2.

### Statistical analysis

The sample size of our trial design meets the minimum safety requirement of 3,000 study participants for the vaccine group, as recommended by the FDA, and WHO guidance.^10,11^ The immunogenicity data are GMT, GMT ratio, and SCR, which are presented with two-sided 95% Confidence Interval (CI). GMT ratio is defined as geometric mean of fold increase of post-study intervention titres over the baseline titres. SCR is defined as the percentage of study subjects with ≥4-fold increase in titres from the baseline or from half of the lower limit of detection (LoD) if baseline is undetectable.

The Safety Set comprises all randomised participants who received at least one dose of study intervention. The immunogenicity set comprises participants who received planned doses of study intervention, had valid immunogenicity results on Day 57, and did not have a major protocol deviation up to Day 57. The PPI Subset was the primary analysis population for the primary and secondary immunogenicity endpoints in this interim analysis.

Participants from different study sites were pooled for statistical analysis. Significance tests (two-tailed, alpha = 0·05) were performed as follows: two-sample t-test or Wilcoxon rank sum test was used for pairwise comparison of GMT and GMT ratios; Pearson’s Chi-square test or Fisher’s exact test (in the case of small cell count or expected count less than five) was used for calculating significance of SCR results; P-value was rounded to four decimal places, if applicable. All statistical analyses were performed using SAS® 9.4 or newer versions (SAS Institute, Cary, NC, USA).

## Results

From 30 Dec 2020 to 2 Apr 2021, 3,844 participants who received at least one dose of study intervention (3,295 received MVC-COV1901 and 549 received placebo) were included in the Safety Set (Figure 1). A total of 1,053 participants fulfilled the definition of the PPI Subset. The mean age of the Safety Set was 45 years (min 20 years, max 89 years), and the male to female percentage was 56·3% to 43·7%. Only one participant was non-Asian (white). Most participants had BMI < 30 kg/m^2^ (89·4%), and negative serology tests for HBsAg, anti-HCV antibody, and HIV antibody at the baseline. A total of 634 (16·5%) participants reported to have comorbidity, most of whom had higher levels of HbA1c than the normal range (13·4%), followed by cardiovascular disease (2·6%) and malignancy (1·1%). Demographic characteristics of the Safety Set are summarised in Table 1.

**Figure 1.**
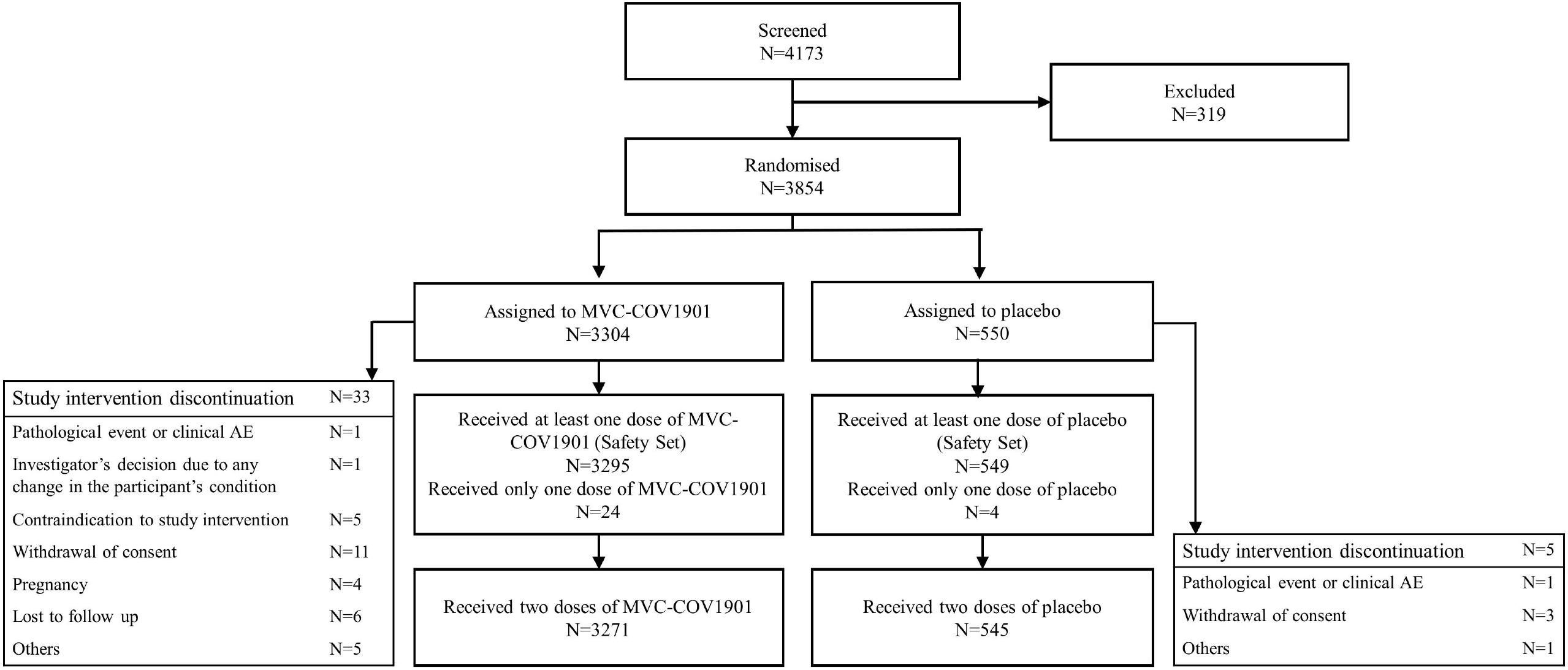
Consort Flow Diagram of the study.

### Safety

The solicited adverse events are summarised in Figure 2 and Supplementary Tables S3 to S5. Overall, 2,510 (65·3%) participants reported solicited local AEs after any dose of the study intervention. The MVC-COV1901 group (2,381 [72·3%]) outnumbered the placebo group (129 [23·5%]) in reporting solicited local AEs, which were mostly mild (Grade 1) to moderate (Grade 2) in severity. After any dose of MVC-COV1901, the most common solicited local AE was pain at the injection site (2,346 [71·2%]), while the most common solicited systemic AE was malaise/fatigue (1,186 [36·0%]). Fever was rarely reported (<1%). Most of the solicited AEs were self-limited in seven days, with a mean duration of less than three days. The occurrence of solicited systemic AEs after any dose of the study intervention in the ≥ 65 years age group (38·7%) was slightly lower than that in the overall Safety Set. Twenty-eight percent of participants (1,081/3,844) reported unsolicited AEs (Supplementary Table S6). The percentages of all participants who reported unsolicited AEs and other AEs in the overall Safety Set and by age group were comparable between the MVC-COV1901 and placebo groups.

**Figure 2.**
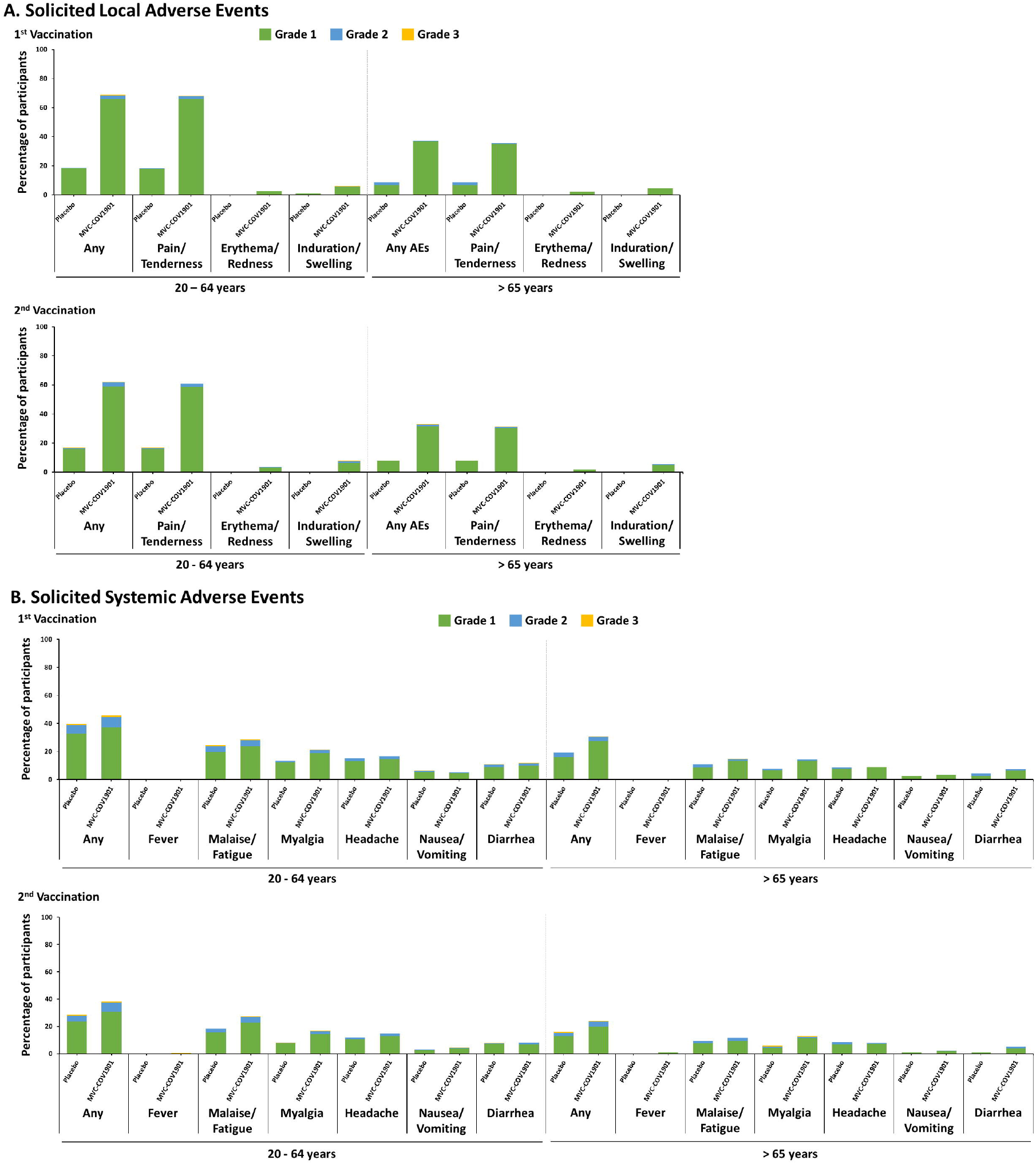
Solicited (A) Local and (B) Systemic Adverse Events within 7 days of first or second dose of study intervention. Severity gradings are as follows: Grade 1 = Mild, Grade 2 = Moderate, Grade 3 = Severe

None of the SAEs were related to the study intervention. One AESI was reported in one participant (< 0·1%) from the MVC-COV1901 group (temporary facial paralysis, possibly related). Neither death nor VAED were reported.

### Immunogenicity

For the PPI Subset of the vaccinated group on D57, the SARS-CoV-2 wild type neutralising antibody GMT was 662·3 (95% CI [628·7, 697·7], and the GMT ratio was 163·2 (95% CI [155·0, 171·9], respectively. Both data were significantly higher (p < 0·0001) than that of the placebo group (Figure 3 and Supplementary Table S7). In vaccinated group, the GMT was higher in the younger adult group 732·9 (95% CI [692·4, 775·7]) than the older adult group 484·5 (95% CI [433·2, 542·0]). The GMTs of NIBSC 20/136, 20/148, 20/144, and 20/140 were 1,620, 258, 181, and 87, respectively. The GMTs of the neutralising antibody on Day 57 for all participants in the PPI Subset, for the younger adult group, and for the older adult group when converted to WHO International Unit (IU) were: 408 IU/mL, 454 IU/mL, and 296 IU/mL, respectively (Figure 3).

**Figure 3.**
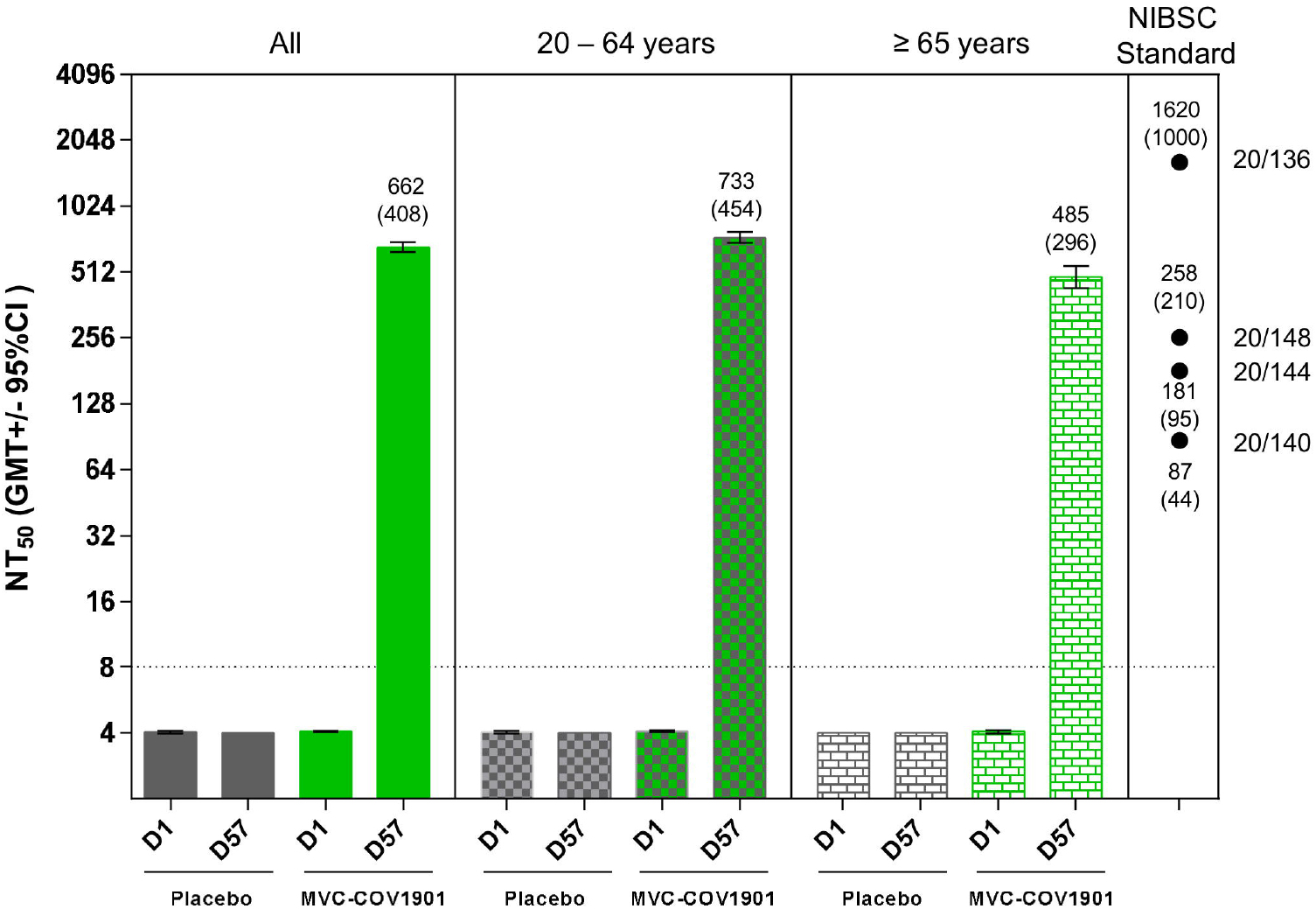
Neutralising antibody titres against live wild type SARS-CoV-2 virus on Day 1 and Day 57. The height of the bar represents GMT, and the error bars represent 95% CI of the GMT. GMT values are marked above the bars that have values over LoD (> 8). The mean value of the triplicated measurement of each NIBSC standard is indicated above or below a dot, and the name of the standard is indicated on the side of the axis. The values in IU/mL are in parentheses.

In the MVC-COV1901 group, the SCR on Day 57 based on the wild type SARS-CoV-2 GMT was 99·8%, with only two participants failing to seroconvert. In the placebo group, no seroconversion was observed (Supplementary Table S8). In both age groups, almost all participants achieved seroconversion (99·9% in younger adults and 99·5% in older adults).

After the first dose (Day 29), the anti-Spike IgG GMT of the PPI Subset increased to 430·5 (95% CI [398·7, 464·8] in the MVC-COV1901 group, whereas it remained at the baseline level in the placebo group. The anti-Spike IgG titre continued to increase to 8,262·2 (95% CI [7,801·9, 8,749·5]) on Day 43 and remained as high as 5745·4 (95% CI [5,464·5, 6,040·6]) on Day 57 (Figure 4 and Supplementary Table S9). The younger adult group had higher anti-Spike IgG GMTs than those of older adult group. The GMTs for the younger adult group were 524 (95% CI [482·7, 569·4]) on Day 29, 9,638 (95% CI [9,070·7, 10,239·9]) on Day 43, and 6,521 (95% CI [6,188·3, 6,871·3]) on Day 57. The GMTs for the older adult group were 233 (95% CI [198·9, 273·1]) on Day 29, 5,126 (95% CI [4,538·3, 5,789·2]) on Day 43, and 3,887 (95% CI [3,476·1, 4,346·7]) on Day 57 (Supplementary Table S9). The anti-Spike IgG GMT on Day 57 for all the participants in the PPI Subset, for the younger adult group, for the older adult group when converted to WHO’s Binding Antibody Unit (BAU/mL) were: 524 BAU/mL, 595 BAU/mL, and 355 BAU/mL, respectively. Supplementary Table S10 illustrates D57 neutralising and binding antibody GMTs converted to WHO’s IU & BAU.

**Figure 4.**
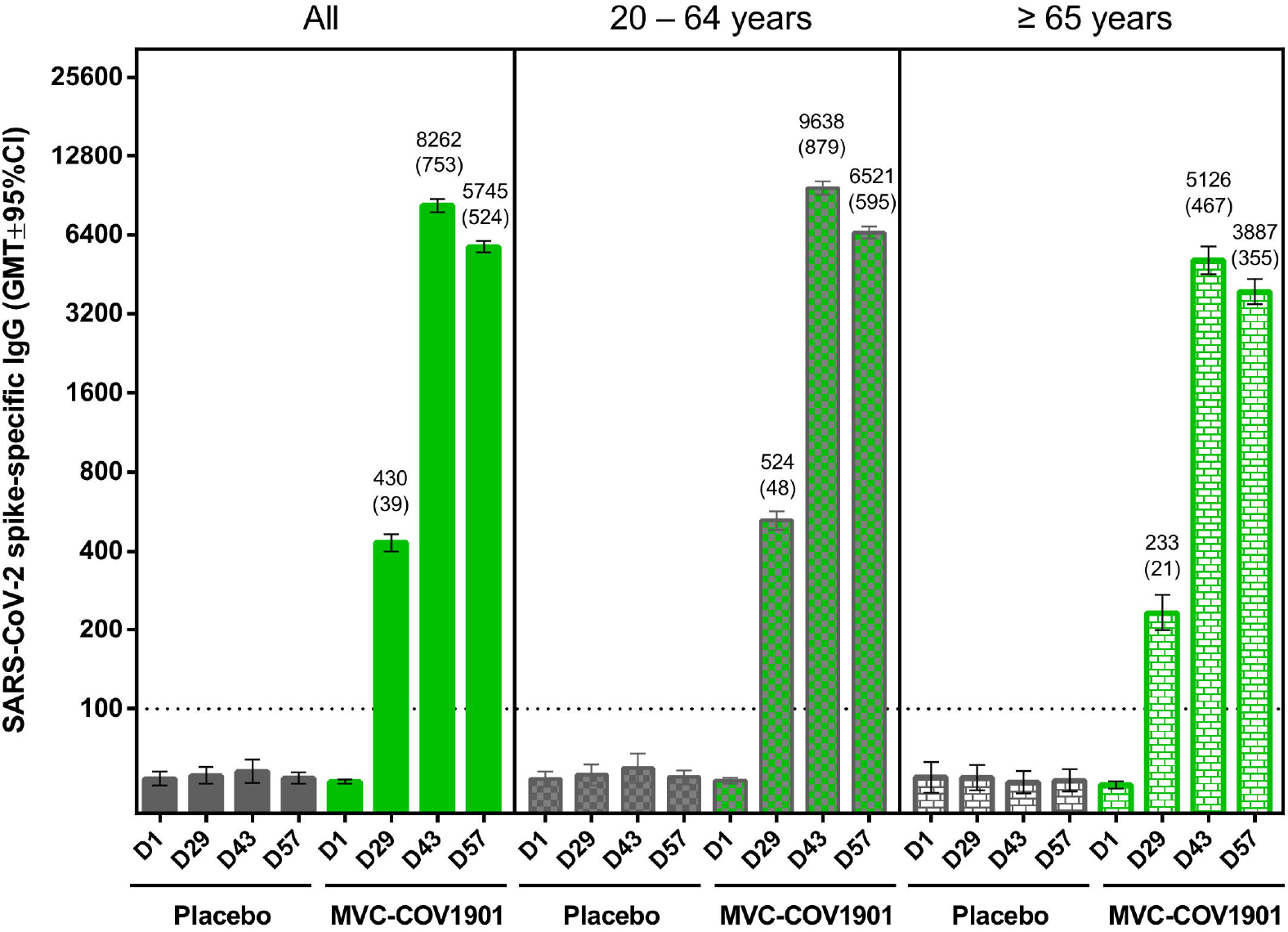
Anti-Spike IgG GMTs measured by ELISA using S-2P as coated antigen on Day 1, Day 29, Day 43, and Day 57. The height of the bar represents GMT, and the error bars represent 95% CI of the GMT. GMT values are marked above the bars that have values above LoD (> 100). The values in BAU/mL are in parentheses.

The SCR of anti-Spike IgG titre was 74·4% in all PPI Subset on Day 29, and it increased to 99·7% on Day 43, and 99·6% on Day 57 (Supplementary Table S11). On Day 29, the SCR was 53·9% in the older adult group, and 80·9% in the younger adult group. However, the SCR for both age groups became comparable on Day 43 (99·5% vs. 99·7%).

## Discussion

This phase 2 trial design followed the principle of the October 2020 US FDA guidance for industry on EUA for vaccines to prevent COVID-19.^10^ By the time of this interim analysis, 3,295 participants have received at least one dose of MVC-COV1901. The median follow-up has reached 63 days, and all study participants have received follow-ups for over a month. The number of participants in the vaccine group (>3,000) and the accumulated safety data published here meet the principles outlined by the US FDA guidance.

The antigen of MVC-COV1901 was designed by Dr. McLellan and Dr. Graham. This antigen design involves a range of molecular modifications of the S-2P prefusion spike protein, furitin cleavage site mutation, and T4 fibritin for trimerization.^4^ The combination of CpG 1018 and aluminium hydroxide with this S-2P prefusion spike protein shows promising enhancements of both T-cell and B-cell immunity, as demonstrated in our phase 1 trial.^7^ The interim analysis of MVC-COV1901 shows that in participants aged 20 years and older, receiving two doses of vaccination is safe, well-tolerated, and reaches favourable levels of neutralising antibody immunogenicity. One of the most distinct findings in our safety profile is the extremely low incidence of fever: 0·3% for the vaccinated group and 0·2% for the placebo group. Compared to other vaccines that have received EUA,^12-14^ MVC-COV1901 causes significantly fewer febrile reactions and has good reactogenicity profile, as seen in this study and our phase 1 trial.^7^

This trial began in December 2020, when SARS-CoV-2 was not endemic in Taiwan. Therefore, very low levels of wildtype neutralising antibody GMTs were detected in all study participants at the baseline (4·06). However, on Day 57 whereas the GMT for the placebo group stayed at 4·0, the GMT for the MVC-COV1901 group spiked to 662·31, which is most likely the results of vaccination.

David Khoury et al. demonstrated that comparing neutralising antibody titres to human convalescent sera (HCS) can be used to predict clinical efficacy for vaccines across different technology platforms.^15^ However, the neutralising antibody titres of convalescent sera vary significantly depending on the degree of disease severity and individual immune response. This variation makes back-to-back comparison of different HCS difficult, if not impossible. To facilitate comparison and avoid mis-extrapolation of the GMT titres on prediction of efficacy, we used two different sets of HCS as parameters to estimate the ratio of our neutralising antibody GMT against the GMT of HCS.

Our first parameter is based on Khoury’s article, which reported that 20% of HCS (54 IU/mL) can approximate 50% vaccine efficacy. Therefore, when we set 270 IU/mL as the GMT of HCS (54 IU/20% = 270 IU/mL), the GMT on Day 57 for the MVC-COV1901 group was 1·51 times that of HCS. The second parameter was based on WHO’s reference panel NIBSC 20/148, which measured at 210 IU/mL in Figure 3. At 210 IU/mL as mid HCS, the GMT on Day 57 for MVC-COV1901 group is 1·94 times that of HCS. When compared to the HCS for both parameters, our neutralising antibody GMTs for all participants in the PPI Subset on Day 57 are approximately 1·51-1·94 times higher.

Another method to predict clinical efficacy is the Binding Antibody Unit (BAU) conversion model. Researchers of the University of Oxford^16^ published the correlates of protection (CoP) for AstraZeneca COVID-19 vaccine against symptomatic and asymptomatic SARS-CoV-2 infection, which is the only CoP published to date. When converted to WHO’s BAU, the report showed that if the anti-Spike IgG level is between 264 and 899, the predicted vaccine efficacy is between 80% to 90%. In our study for MVC-COV1901, the anti-Spike IgG GMT on Day 57 for all PPI Subset was 524 BAU/mL; for the younger adult group was 595 BAU/mL; for the older adult group was 355 BAU/mL. Even though the effects of cellular immunity and ethnic differences remain unclear, in both the IU and the BAU conversion models, MVC-COV1901 has yielded comparable vaccine efficacy predictions.

For regulatory authorities to use CoP for cross-vaccine comparison to predict clinical efficacy, a standardised assay which converts GMT to IU or BAU will be a pre-requisite.^17,18^ This conversion assay would expedite vaccine approval, facilitate cross-checking of immunogenicity between trials, and enable data comparisons among different labs. This study is, to our knowledge, the first to report neutralising antibody titres and anti-Spike IgG titres using WHO’s IU and BAU for a COVID-19 vaccine in phase 2. We applied the IU conversion methodology to validate the consistency of immunogenicity between our phase 1 and phase 2 trials from different labs. Using WHO’s IU, our phase 1 GMT on Day 57 was converted from 52·2 to 351·2 IU/mL, and our phase 2 GMT on Day 57 was converted from 663·2 to 408 IU/mL. The converted GMTs indicate that the immunogenicity data from our phase 1 and phase 2 trials is comparable. Our results demonstrate the many benefits of standardised assays which may facilitate the application of CoP for regulatory authorities to consider.

It is paramount for a COVID-19 vaccine to render protection for people at higher risk of severe complications. Several reports show that elderly patients with chronic comorbidities have higher mortality rates of COVID-19.^19,20^ In this phase 2 trial, we included older adult participants with various chronic comorbidities in stable conditions, which allowed us to assess the safety and immunogenicity profiles of MVC-COV1901 for this age group. Since the safety profiles, neutralising titres, tolerability, and vaccine-related SAEs are comparable between the younger adult and older adult groups, MVC-COV1901 is suitable for older adults over 65 years of age with comorbidities.

We acknowledge the limitations of our current study. The low viral transmission rate in Taiwan at the time of the study, coupled with the relatively small size of the placebo group, hindered observation of both vaccine efficacy as the exploratory endpoint and the risk of VAED. The short duration of the follow-up period before the interim analysis prevented us from assessing the durability of immune responses after Day 57; the long-term follow-up will be available upon completion of this phase 2 trial in October 2021. The racial diversity of study participants is restricted to ethnicities in Taiwan. These limitations will be addressed in our phase 3 trials, which will be conducted in regions outside Taiwan where COVID-19 infection rates are high.

To conclude, this interim analysis shows that MVC-COV1901 has good safety profiles and promising neutralising antibody titres. MVC-COV1901 is safe, well-tolerated, and rarely causes febrile reactions in both young adults and older adults. MVC-COV1901 induces high neutralising antibody and anti-Spike IgG GMTs, and its seroconversion rate is nearly 100% on Day 57. Using WHO’s IU and BAU conversion models, the predicted clinical efficacy for MVC-COV1901 is comparable for both methods. The phase 2 trial results support the advancement of MVC-COV1901 in subsequent phase 3 trials. MVC-COV1901 has recently been granted EUA in Taiwan.

## Supporting information

Supplementary appedix

## Data Availability

Data are available upon reasonable request

## Author contributions

Hsieh SM, Lin TY, Lien CE and Tai IC had full access to all the data in this study and take responsibility for the integrity of the data and the accuracy of the data analysis. Lin TY and Tai IC contributed equally and are joint corresponding authors.

Concept and design: Hsieh SM and Tai IC.

Acquisition or interpretation of data: Hsieh SM, Liu MC, Chen YS, Lee WS, Hwang SJ, Cheng SH, Hwang KP, Ko WC, Wang NC, and Lee YL.

Drafting and preparation of the manuscript: Hsieh SM, Liu Mc, Chen YS, Lee WS, Hwang SJ, Cheng SH, Lien CE, Tai IC and Lin TY.

Laboratory assays set up, assay conduction, and data analysis: Lin YL, Huang CG, Shih SR, Liao CC, Liang JJ, Chang CS.

Administrative, technical, and material support: Chen C.

All authors reviewed and approved the final version of the manuscript.

## Acknowledgements

The authors would like to thank:

Dr. Stanley Chang, Dr. Meei-Yun Lin, and Dr. Luke Tzu-Chi Liu at Medigen Vaccine Biologics Corp. for manuscript drafting, editing, and revision.

All the participants for their dedication to this trial.

The investigational staff at National Taiwan University Hospital, Taiwan Taipei Veterans General Hospital, Tri-Service General Hospital, Taipei Veterans General Hospital, Taipei Medical University Hospital, Taipei Municipal Wan Fang Hospital, Linkou Chang Gung Medical Hospital, Tao-Yuan General Hospital, China Medical University Hospital, Changhua Christian Hospital, National Cheng Kung University Hospital, and Kaohsiung Medical University Chung-Ho Memorial Hospital, Clinipace Clinical Research Team, Taiwan., and Hao-Yuan Cheng, Criss Cheng, Meng-Ju Tsai at Medigen Vaccine Biologics Corp. for the conduction of the trial.

Dr. Barney S. Graham, at Vaccine Research Centre, National Institute of Allergy and Infectious Diseases, U.S.A. for the development of S-2P prefusion protein.

Dr. Tsun□ Yung Kuo and team at Department of Biotechnology and Animal Science, National Ilan University, Ilan, Taiwan for technical guidance and scientific advice.

Dr. Robert Janssen at Dynavax Technologies for providing critical intellectual content of the manuscript.

Dynavax Technologies for providing the CpG 1018 adjuvant.

Dr. Wei-Cheng Lian, Dr. Erh-Fang Hsieh, Dr. Yi-Jiun Li at Medigen Vaccine Biologics Corp. for collaboration on vaccine production, significant contribution to the Investigational New Drug (IND) application, and laboratory assay development.

The members of the Independent Data Monitoring Committee.

Team members at Protech Pharmaservices Corp. for spike specific IgG ELISA assay.

Team members at the Department of Laboratory Medicine, Linkou Chang Gung Memorial Hospital, and Institute of Biomedical Sciences, Academia Sinica for neutralisation assay conduction.

