## Supplementary appedix for "Safety and Immunogenicity of CpG 1018 and Aluminium Hydroxide-Adjuvanted SARS-CoV-2 S-2P Protein Vaccine MVC-COV1901: A Large-Scale Double-Blind, Randomised, Placebo-Controlled Phase 2 Trial"

Supplementary Appendix

Supplementary Tables

Table S1. Solicited Adverse Events after First Dosing

|  | All, n (%) | | | 20 to 64 years, n (%) | | | ≥ 65 years, n (%) | | |
| --- | --- | --- | --- | --- | --- | --- | --- | --- | --- |
|  | MVC-COV1901  (N = 3295) | Placebo  (N = 549) | Total  (N = 3844) | MVC-COV1901  (N = 2575) | Placebo  (N = 431) | Total  (N = 3006) | MVC-COV1901  (N = 720) | Placebo  (N = 118) | Total  (N = 838) |
| Any Solicited Local AEs | **2040 (61.9)** | **90 (16.4)** | **2130 (55.4)** | **1772 (68.8)** | **80 (18.6)** | **1852 (61.6)** | **268 (37.2)** | **10 (8.5)** | **278 (33.2)** |
| **Pain/Tenderness** | **2011 (61.0)** | **88 (16.0)** | **2099 (54.6)** | **1755 (68.2)** | **78 (18.1)** | **1833 (61.0)** | **256 (35.6)** | **10 (8.5)** | **266 (31.7)** |
| Grade 1 | 1952 (59.2) | 85 (15.5) | 2037 (53.0) | 1699 (66.0) | 77 (17.9) | 1776 (59.1) | 253 (35.1) | 8 (6.8) | 261 (31.1) |
| Grade 2 | 51 (1.5) | 3 (0.5) | 54 (1.4) | 48 (1.9) | 1 (0.2) | 49 (1.6) | 3 (0.4) | 2 (1.7) | 5 (0.6) |
| Grade 3 | 8 (0.2) | 0 | 8 (0.2) | 8 (0.3) | 0 | 8 (0.3) | 0 | 0 | 0 |
| **Induration/Swelling** | **187 (5.7)** | **4 (0.7)** | **191 (5.0)** | **154 (6.0)** | **4 (0.9)** | **158 (5.3)** | **33 (4.6)** | **0** | **33 (3.9)** |
| Grade 1 | 171 (5.2) | 4 (0.7) | 175 (4.6) | 138 (5.4) | 4 (0.9) | 142 (4.7) | 33 (4.6) | 0 | 33 (3.9) |
| Grade 2 | 15 (0.5) | 0 | 15 (0.4) | 15 (0.6) | 0 | 15 (0.5) | 0 | 0 | 0 |
| Grade 3 | 1 (<0.1) | 0 | 1 (<0.1) | 1 (<0.1) | 0 | 1 (<0.1) | 0 | 0 | 0 |
| **Erythema/Redness** | **80 (2.4)** | **0** | **80 (2.1)** | **66 (2.6)** | **0** | **66 (2.2)** | **14 (1.9)** | **0** | **14 (1.7)** |
| Grade 1 | 78 (2.4) | 0 | 78 (2.0) | 64 (2.5) | 0 | 64 (2.1) | 14 (1.9) | 0 | 14 (1.7) |
| Grade 2 | 2 (0.1) | 0 | 2 (0.1) | 2 (0.1) | 0 | 2 (0.1) | 0 | 0 | 0 |
| Any Solicited Systemic AEs | **1400 (42.5)** | **194 (35.3)** | **1594 (41.5)** | **1179 (45.8)** | **171 (39.7)** | **1350 (44.9)** | **221 (30.7)** | **23 (19.5)** | **244 (29.1)** |
| **Malaise/Fatigue** | **842 (25.6)** | **118 (21.5)** | **960 (25.0)** | **736 (28.6)** | **105 (24.4)** | **841 (28.0)** | **106 (14.7)** | **13 (11.0)** | **119 (14.2)** |
| Grade 1 | 716 (21.7) | 95 (17.3) | 811 (21.1) | 619 (24.0) | 85 (19.7) | 704 (23.4) | 97 (13.5) | 10 (8.5) | 107 (12.8) |
| Grade 2 | 114 (3.5) | 21 (3.8) | 135 (3.5) | 106 (4.1) | 18 (4.2) | 124 (4.1) | 8 (1.1) | 3 (2.5) | 11 (1.3) |
| Grade 3 | 12 (0.4) | 2 (0.4) | 14 (0.4) | 11 (0.4) | 2 (0.5) | 13 (0.4) | 1 (0.1) | 0 | 1 (0.1) |
| **Myalgia** | **655 (19.9)** | **66 (12.0)** | **721 (18.8)** | **552 (21.4)** | **57 (13.2)** | **609 (20.3)** | **103 (14.3)** | **9 (7.6)** | **112 (13.4)** |
| Grade 1 | 574 (17.4) | 62 (11.3) | 636 (16.5) | 478 (18.6) | 54 (12.5) | 532 (17.7) | 96 (13.3) | 8 (6.8) | 104 (12.4) |
| Grade 2 | 72 (2.2) | 4 (0.7) | 76 (2.0) | 65 (2.5) | 3 (0.7) | 68 (2.3) | 7 (1.0) | 1 (0.8) | 8 (1.0) |
| Grade 3 | 9 (0.3) | 0 | 9 (0.2) | 9 (0.3) | 0 | 9 (0.3) | 0 | 0 | 0 |
| **Headache** | **496 (15.1)** | **75 (13.7)** | **571 (14.9)** | **432 (16.8)** | **65 (15.1)** | **497 (16.5)** | **64 (8.9)** | **10 (8.5)** | **74 (8.8)** |
| Grade 1 | 442 (13.4) | 65 (11.8) | 507 (13.2) | 379 (14.7) | 56 (13.0) | 435 (14.5) | 63 (8.8) | 9 (7.6) | 72 (8.6) |
| Grade 2 | 49 (1.5) | 10 (1.8) | 59 (1.5) | 48 (1.9) | 9 (2.1) | 57 (1.9) | 1 (0.1) | 1 (0.8) | 2 (0.2) |
| Grade 3 | 5 (0.2) | 0 | 5 (0.1) | 5 (0.2) | 0 | 5 (0.2) | 0 | 0 | 0 |
| **Diarrhea** | **355 (10.8)** | **52 (9.5)** | **407 (10.6)** | **302 (11.7)** | **47 (10.9)** | **349 (11.6)** | **53 (7.4)** | **5 (4.2)** | **58 (6.9)** |
| Grade 1 | 309 (9.4) | 41 (7.5) | 350 (9.1) | 263 (10.2) | 38 (8.8) | 301 (10.0) | 46 (6.4) | 3 (2.5) | 49 (5.8) |
| Grade 2 | 39 (1.2) | 10 (1.8) | 49 (1.3) | 32 (1.2) | 8 (1.9) | 40 (1.3) | 7 (1.0) | 2 (1.7) | 9 (1.1) |
| Grade 3 | 7 (0.2) | 1 (0.2) | 8 (0.2) | 7 (0.3) | 1 (0.2) | 8 (0.3) | 0 | 0 | 0 |
| **Nausea/Vomiting** | **159 (4.8)** | **30 (5.5)** | **189 (4.9)** | **135 (5.2)** | **27 (6.3)** | **162 (5.4)** | **24 (3.3)** | **3 (2.5)** | **27 (3.2)** |
| Grade 1 | 143 (4.3) | 27 (4.9) | 170 (4.4) | 119 (4.6) | 24 (5.6) | 143 (4.8) | 24 (3.3) | 3 (2.5) | 27 (3.2) |
| Grade 2 | 15 (0.5) | 2 (0.4) | 17 (0.4) | 15 (0.6) | 2 (0.5) | 17 (0.6) | 0 | 0 | 0 |
| Grade 3 | 1 (<0.1) | 1 (0.2) | 2 (0.1) | 1 (<0.1) | 1 (0.2) | 2 (0.1) | 0 | 0 | 0 |
| **Fever** | **13 (0.4)** | **1 (0.2)** | **14 (0.4)** | **11 (0.4)** | **1 (0.2)** | **12 (0.4)** | **2 (0.3)** | **0** | **2 (0.2)** |
| Grade 1 | 8 (0.2) | 1 (0.2) | 9 (0.2) | 7 (0.3) | 1 (0.2) | 8 (0.3) | 1 (0.1) | 0 | 1 (0.1) |
| Grade 2 | 3 (0.1) | 0 | 3 (0.1) | 3 (0.1) | 0 | 3 (0.1) | 0 | 0 | 0 |
| Grade 3 | 1 (<0.1) | 0 | 1 (<0.1) | 1 (<0.1) | 0 | 1 (<0.1) | 0 | 0 | 0 |
| Grade4 | 1 (<0.1) | 0 | 1 (<0.1) | 0 | 0 | 0 | 1 (0.1) | 0 | 1 (0.1) |

Table S2. Solicited Adverse Events after Second Dosing

|  | All, n (%) | | | 20 to 64 years, n (%) | | | ≥ 65 years, n (%) | | |
| --- | --- | --- | --- | --- | --- | --- | --- | --- | --- |
|  | MVC-COV1901  (N = 3295) | Placebo  (N = 549) | Total  (N = 3844) | MVC-COV1901  (N = 2575) | Placebo  (N = 431) | Total  (N = 3006) | MVC-COV1901  (N = 720) | Placebo  (N = 118) | Total  (N = 838) |
| Any Solicited Local AEs | **1832 (55.6)** | **81 (14.8)** | **1913 (49.8)** | **1595 (61.9)** | **72 (16.7)** | **1667 (55.5)** | **237 (32.9)** | **9 (7.6)** | **246 (29.4)** |
| **Pain/Tenderness** | **1791 (54.4)** | **81 (14.8)** | **1872 (48.7)** | **1566 (60.8)** | **72 (16.7)** | **1638 (54.5)** | **225 (31.3)** | **9 (7.6)** | **234 (27.9)** |
| Grade 1 | 1727 (52.4) | 78 (14.2) | 1805 (47.0) | 1508 (58.6) | 69 (16.0) | 1577 (52.5) | 219 (30.4) | 9 (7.6) | 228 (27.2) |
| Grade 2 | 59 (1.8) | 2 (0.4) | 61 (1.6) | 54 (2.1) | 2 (0.5) | 56 (1.9) | 5 (0.7) | 0 | 5 (0.6) |
| Grade 3 | 5 (0.2) | 1 (0.2) | 6 (0.2) | 4 (0.2) | 1 (0.2) | 5 (0.2) | 1 (0.1) | 0 | 1 (0.1) |
| **Induration/Swelling** | **233 (7.1)** | **1 (0.2)** | **234 (6.1)** | **194 (7.5)** | **1 (0.2)** | **195 (6.5)** | **39 (5.4)** | **0** | **39 (4.7)** |
| Grade 1 | 200 (6.1) | 1 (0.2) | 201 (5.2) | 165 (6.4) | 1 (0.2) | 166 (5.5) | 35 (4.9) | 0 | 35 (4.2) |
| Grade 2 | 32 (1.0) | 0 | 32 (0.8) | 28 (1.1) | 0 | 28 (0.9) | 4 (0.6) | 0 | 4 (0.5) |
| Grade 3 | 1 (<0.1) | 0 | 1 (<0.1) | 1 (<0.1) | 0 | 1 (<0.1) | 0 | 0 | 0 |
| **Erythema/Redness** | **98 (3.0)** | **0** | **98 (2.5)** | **85 (3.3)** | **0** | **85 (2.8)** | **13 (1.8)** | **0** | **13 (1.6)** |
| Grade 1 | 94 (2.9) | 0 | 94 (2.4) | 81 (3.1) | 0 | 81 (2.7) | 13 (1.8) | 0 | 13 (1.6) |
| Grade 2 | 4 (0.1) | 0 | 4 (0.1) | 4 (0.2) | 0 | 4 (0.1) | 0 | 0 | 0 |
| Any Solicited Systemic AEs | **1159 (35.2)** | **142 (25.9)** | **1301 (33.8)** | **986 (38.3)** | **123 (28.5)** | **1109 (36.9)** | **173 (24.0)** | **19 (16.1)** | **192 (22.9)** |
| **Malaise/Fatigue** | **788 (23.9)** | **91 (16.6)** | **879 (22.9)** | **705 (27.4)** | **80 (18.6)** | **785 (26.1)** | **83 (11.5)** | **11 (9.3)** | **94 (11.2)** |
| Grade 1 | 656 (19.9) | 76 (13.8) | 732 (19.0) | 587 (22.8) | 67 (15.5) | 654 (21.8) | 69 (9.6) | 9 (7.6) | 78 (9.3) |
| Grade 2 | 121 (3.7) | 14 (2.6) | 135 (3.5) | 108 (4.2) | 12 (2.8) | 120 (4.0) | 13 (1.8) | 2 (1.7) | 15 (1.8) |
| Grade 3 | 11 (0.3) | 1 (0.2) | 12 (0.3) | 10 (0.4) | 1 (0.2) | 11 (0.4) | 1 (0.1) | 0 | 1 (0.1) |
| **Myalgia** | **521 (15.8)** | **43 (7.8)** | **564 (14.7)** | **430 (16.7)** | **36 (8.4)** | **466 (15.5)** | **91 (12.6)** | **7 (5.9)** | **98 (11.7)** |
| Grade 1 | 449 (13.6) | 38 (6.9) | 487 (12.7) | 368 (14.3) | 33 (7.7) | 401 (13.3) | 81 (11.3) | 5 (4.2) | 86 (10.3) |
| Grade 2 | 63 (1.9) | 2 (0.4) | 65 (1.7) | 55 (2.1) | 1 (0.2) | 56 (1.9) | 8 (1.1) | 1 (0.8) | 9 (1.1) |
| Grade 3 | 9 (0.3) | 3 (0.5) | 12 (0.3) | 7 (0.3) | 2 (0.5) | 9 (0.3) | 2 (0.3) | 1 (0.8) | 3 (0.4) |
| **Headache** | **435 (13.2)** | **61 (11.1)** | **496 (12.9)** | **378 (14.7)** | **51 (11.8)** | **429 (14.3)** | **57 (7.9)** | **10 (8.5)** | **67 (8.0)** |
| Grade 1 | 378 (11.5) | 54 (9.8) | 432 (11.2) | 326 (12.7) | 46 (10.7) | 372 (12.4) | 52 (7.2) | 8 (6.8) | 60 (7.2) |
| Grade 2 | 55 (1.7) | 7 (1.3) | 62 (1.6) | 50 (1.9) | 5 (1.2) | 55 (1.8) | 5 (0.7) | 2 (1.7) | 7 (0.8) |
| Grade 3 | 2 (0.1) | 0 | 2 (0.1) | 2 (0.1) | 0 | 2 (0.1) | 0 | 0 | 0 |
| **Diarrhea** | **248 (7.5)** | **35 (6.4)** | **283 (7.4)** | **211 (8.2)** | **34 (7.9)** | **245 (8.2)** | **37 (5.1)** | **1 (0.8)** | **38 (4.5)** |
| Grade 1 | 202 (6.1) | 32 (5.8) | 234 (6.1) | 174 (6.8) | 31 (7.2) | 205 (6.8) | 28 (3.9) | 1 (0.8) | 29 (3.5) |
| Grade 2 | 42 (1.3) | 2 (0.4) | 44 (1.1) | 34 (1.3) | 2 (0.5) | 36 (1.2) | 8 (1.1) | 0 | 8 (1.0) |
| Grade 3 | 4 (0.1) | 1 (0.2) | 5 (0.1) | 3 (0.1) | 1 (0.2) | 4 (0.1) | 1 (0.1) | 0 | 1 (0.1) |
| **Nausea/Vomiting** | **126 (3.8)** | **13 (2.4)** | **139 (3.6)** | **110 (4.3)** | **12 (2.8)** | **122 (4.1)** | **16 (2.2)** | **1 (0.8)** | **17 (2.0)** |
| Grade 1 | 112 (3.4) | 12 (2.2) | 124 (3.2) | 97 (3.8) | 11 (2.6) | 108 (3.6) | 15 (2.1) | 1 (0.8) | 16 (1.9) |
| Grade 2 | 13 (0.4) | 1 (0.2) | 14 (0.4) | 12 (0.5) | 1 (0.2) | 13 (0.4) | 1 (0.1) | 0 | 1 (0.1) |
| Grade 3 | 1 (<0.1) | 0 | 1 (<0.1) | 1 (<0.1) | 0 | 1 (<0.1) | 0 | 0 | 0 |
| **Fever** | **10 (0.3)** | **1 (0.2)** | **11 (0.3)** | **5 (0.2)** | **1 (0.2)** | **6 (0.2)** | **5 (0.7)** | **0** | **5 (0.6)** |
| Grade 1 | 6 (0.2) | 1 (0.2) | 7 (0.2) | 1 (<0.1) | 1 (0.2) | 2 (0.1) | 5 (0.7) | 0 | 5 (0.6) |
| Grade 2 | 3 (0.1) | 0 | 3 (0.1) | 3 (0.1) | 0 | 3 (0.1) | 0 | 0 | 0 |
| Grade 3 | 1 (<0.1) | 0 | 1 (<0.1) | 1 (<0.1) | 0 | 1 (<0.1) | 0 | 0 | 0 |

Table S3. Solicited Adverse Events After any Dosing

|  | All, n (%) | | | 20 to 64 years, n (%) | | | ≥ 65 years, n (%) | | |
| --- | --- | --- | --- | --- | --- | --- | --- | --- | --- |
|  | MVC-COV1901  (N = 3295) | Placebo  (N = 549) | Total  (N = 3844) | MVC-COV1901  (N = 2575) | Placebo  (N = 431) | Total  (N = 3006) | MVC-COV1901  (N = 720) | Placebo  (N = 118) | Total  (N = 838) |
| Any Solicited Local AEs | **2381 (72.3)** | **129 (23.5)** | **2510 (65.3)** | **2030 (78.8)** | **113 (26.2)** | **2143 (71.3)** | **351 (48.8)** | **16 (13.6)** | **367 (43.8)** |
| **Pain/Tenderness** | **2346 (71.2)** | **128 (23.3)** | **2474 (64.4)** | **2009 (78.0)** | **112 (26.0)** | **2121 (70.6)** | **337 (46.8)** | **16 (13.6)** | **353 (42.1)** |
| Grade 1 | 2237 (67.9) | 123 (22.4) | 2360 (61.4) | 1907 (74.1) | 109 (25.3) | 2016 (67.1) | 330 (45.8) | 14 (11.9) | 344 (41.1) |
| Grade 2 | 97 (2.9) | 4 (0.7) | 101 (2.6) | 91 (3.5) | 2 (0.5) | 93 (3.1) | 6 (0.8) | 2 (1.7) | 8 (1.0) |
| Grade 3 | 12 (0.4) | 1 (0.2) | 13 (0.3) | 11 (0.4) | 1 (0.2) | 12 (0.4) | 1 (0.1) | 0 | 1 (0.1) |
| **Induration/Swelling** | **347 (10.5)** | **5 (0.9)** | **352 (9.2)** | **286 (11.1)** | **5 (1.2)** | **291 (9.7)** | **61 (8.5)** | **0** | **61 (7.3)** |
| Grade 1 | 303 (9.2) | 5 (0.9) | 308 (8.0) | 246 (9.6) | 5 (1.2) | 251 (8.3) | 57 (7.9) | 0 | 57 (6.8) |
| Grade 2 | 42 (1.3) | 0 | 42 (1.1) | 38 (1.5) | 0 | 38 (1.3) | 4 (0.6) | 0 | 4 (0.5) |
| Grade 3 | 2 (0.1) | 0 | 2 (0.1) | 2 (0.1) | 0 | 2 (0.1) | 0 | 0 | 0 |
| **Erythema/Redness** | **161 (4.9)** | **0** | **161 (4.2)** | **138 (5.4)** | **0** | **138 (4.6)** | **23 (3.2)** | **0** | **23 (2.7)** |
| Grade 1 | 155 (4.7) | 0 | 155 (4.0) | 132 (5.1) | 0 | 132 (4.4) | 23 (3.2) | 0 | 23 (2.7) |
| Grade 2 | 6 (0.2) | 0 | 6 (0.2) | 6 (0.2) | 0 | 6 (0.2) | 0 | 0 | 0 |
| Any Solicited Systemic AEs | **1774 (53.8)** | **249 (45.4)** | **2023 (52.6)** | **1484 (57.6)** | **215 (49.9)** | **1699 (56.5)** | **290 (40.3)** | **34 (28.8)** | **324 (38.7)** |
| **Malaise/Fatigue** | **1186 (36.0)** | **163 (29.7)** | **1349 (35.1)** | **1036 (40.2)** | **142 (32.9)** | **1178 (39.2)** | **150 (20.8)** | **21 (17.8)** | **171 (20.4)** |
| Grade 1 | 961 (29.2) | 134 (24.4) | 1095 (28.5) | 831 (32.3) | 117 (27.1) | 948 (31.5) | 130 (18.1) | 17 (14.4) | 147 (17.5) |
| Grade 2 | 203 (6.2) | 27 (4.9) | 230 (6.0) | 185 (7.2) | 23 (5.3) | 208 (6.9) | 18 (2.5) | 4 (3.4) | 22 (2.6) |
| Grade 3 | 22 (0.7) | 2 (0.4) | 24 (0.6) | 20 (0.8) | 2 (0.5) | 22 (0.7) | 2 (0.3) | 0 | 2 (0.2) |
| **Myalgia** | **908 (27.6)** | **91 (16.6)** | **999 (26.0)** | **757 (29.4)** | **79 (18.3)** | **836 (27.8)** | **151 (21.0)** | **12 (10.2)** | **163 (19.5)** |
| Grade 1 | 764 (23.2) | 83 (15.1) | 847 (22.0) | 629 (24.4) | 73 (16.9) | 702 (23.4) | 135 (18.8) | 10 (8.5) | 145 (17.3) |
| Grade 2 | 126 (3.8) | 5 (0.9) | 131 (3.4) | 112 (4.3) | 4 (0.9) | 116 (3.9) | 14 (1.9) | 1 (0.8) | 15 (1.8) |
| Grade 3 | 18 (0.5) | 3 (0.5) | 21 (0.5) | 16 (0.6) | 2 (0.5) | 18 (0.6) | 2 (0.3) | 1 (0.8) | 3 (0.4) |
| **Headache** | **730 (22.2)** | **110 (20.0)** | **840 (21.9)** | **631 (24.5)** | **94 (21.8)** | **725 (24.1)** | **99 (13.8)** | **16 (13.6)** | **115 (13.7)** |
| Grade 1 | 630 (19.1) | 95 (17.3) | 725 (18.9) | 537 (20.9) | 81 (18.8) | 618 (20.6) | 93 (12.9) | 14 (11.9) | 107 (12.8) |
| Grade 2 | 93 (2.8) | 15 (2.7) | 108 (2.8) | 87 (3.4) | 13 (3.0) | 100 (3.3) | 6 (0.8) | 2 (1.7) | 8 (1.0) |
| Grade 3 | 7 (0.2) | 0 | 7 (0.2) | 7 (0.3) | 0 | 7 (0.2) | 0 | 0 | 0 |
| **Diarrhea** | **497 (15.1)** | **69 (12.6)** | **566 (14.7)** | **422 (16.4)** | **63 (14.6)** | **485 (16.1)** | **75 (10.4)** | **6 (5.1)** | **81 (9.7)** |
| Grade 1 | 411 (12.5) | 56 (10.2) | 467 (12.1) | 350 (13.6) | 52 (12.1) | 402 (13.4) | 61 (8.5) | 4 (3.4) | 65 (7.8) |
| Grade 2 | 75 (2.3) | 11 (2.0) | 86 (2.2) | 62 (2.4) | 9 (2.1) | 71 (2.4) | 13 (1.8) | 2 (1.7) | 15 (1.8) |
| Grade 3 | 11 (0.3) | 2 (0.4) | 13 (0.3) | 10 (0.4) | 2 (0.5) | 12 (0.4) | 1 (0.1) | 0 | 1 (0.1) |
| **Nausea/Vomiting** | **254 (7.7)** | **37 (6.7)** | **291 (7.6)** | **219 (8.5)** | **33 (7.7)** | **252 (8.4)** | **35 (4.9)** | **4 (3.4)** | **39 (4.7)** |
| Grade 1 | 226 (6.9) | 33 (6.0) | 259 (6.7) | 192 (7.5) | 29 (6.7) | 221 (7.4) | 34 (4.7) | 4 (3.4) | 38 (4.5) |
| Grade 2 | 26 (0.8) | 3 (0.5) | 29 (0.8) | 25 (1.0) | 3 (0.7) | 28 (0.9) | 1 (0.1) | 0 | 1 (0.1) |
| Grade 3 | 2 (0.1) | 1 (0.2) | 3 (0.1) | 2 (0.1) | 1 (0.2) | 3 (0.1) | 0 | 0 | 0 |
| **Fever** | **23 (0.7)** | **2 (0.4)** | **25 (0.7)** | **16 (0.6)** | **2 (0.5)** | **18 (0.6)** | **7 (1.0)** | **0** | **7 (0.8)** |
| Grade 1 | 14 (0.4) | 2 (0.4) | 16 (0.4) | 8 (0.3) | 2 (0.5) | 10 (0.3) | 6 (0.8) | 0 | 6 (0.7) |
| Grade 2 | 6 (0.2) | 0 | 6 (0.2) | 6 (0.2) | 0 | 6 (0.2) | 0 | 0 | 0 |
| Grade 3 | 2 (0.1) | 0 | 2 (0.1) | 2 (0.1) | 0 | 2 (0.1) | 0 | 0 | 0 |
| Grade 4 | 1 (<0.1) | 0 | 1 (<0.1) | 0 | 0 | 0 | 1 (<0.1) | 0 | 1 (<0.1) |

Table S4. Overall Summary of Unsolicited Adverse Events and Other Adverse Events

|  | **All, n (%)** | | | 20 to 64 years, n (%) | | | ≥ 65 years, n (%) | | |
| --- | --- | --- | --- | --- | --- | --- | --- | --- | --- |
|  | MVC-COV1901  **(N = 3295)** | Placebo  **(N = 549)** | Total  **(N = 3844)** | MVC-COV1901  (N = 2575) | Placebo  (N = 431) | Total  (N = 3006) | MVC-COV1901  (N = 720) | Placebo  (N = 118) | Total  (N = 838) |
| Unsolicited AEs | 932 (28.3) | 149 (27.1) | 1081 (28.1) | 767 (29.8) | 123 (28.5) | 890 (29.6) | 165 (22.9) | 26 (22.0) | 191 (22.8) |
| Related unsolicited AEs | 406 (12.3) | 62 (11.3) | 468 (12.2) | 340 (13.2) | 56 (13.0) | 396 (13.2) | 66 (9.2) | 6 (5.1) | 72 (8.6) |
| Unsolicited AEs ≥ Grade 3 | 93 (2.8) | 11 (2.0) | 104 (2.7) | 86 (3.3) | 10 (2.3) | 96 (3.2) | 7 (1.0) | 1 (0.8) | 8 (1.0) |
| Related unsolicited AEs ≥ Grade 3 | 21 (0.6) | 4 (0.7) | 25 (0.7) | 20 (0.8) | 4 (0.9) | 24 (0.8) | 1 (0.1) | 0 | 1 (0.1) |
| SAEs | 18 (0.5) | 1 (0.2) | 19 (0.5) | 16 (0.6) | 1 (0.2) | 17 (0.6) | 2 (0.3) | 0 | 2 (0.2) |
| Related SAEs | 0 | 0 | 0 | 0 | 0 | 0 | 0 | 0 | 0 |
| AESI | 1 (<0.1) | 0 | 1 (<0.1) | 0 | 0 | 0 | 1 (0.1) | 0 | 1 (0.1) |
| VAED | 0 | 0 | 0 | 0 | 0 | 0 | 0 | 0 | 0 |
| AEs leading to study intervention discontinuation | 2 (0.1) | 1 (0.2) | 3 (0.1) | 0 | 1 (0.2) | 1 (<0.1) | 2 (0.3) | 0 | 2 (0.2) |
| AEs leading to study withdrawal | 1 (<0.1) | 0 | 1 (<0.1) | 0 | 0 | 0 | 1 (0.1) | 0 | 1 (0.1) |
| Death | 0 | 0 | 0 | 0 | 0 | 0 | 0 | 0 | 0 |

Abbreviations: AE = adverse event; AESI = adverse events of special interest; CI = confidence interval; N = number of participants in the population; n = number of participants with events; SAE = serious adverse event; VAED = vaccine-associated enhanced disease

Table S5. Wild Type SARS-CoV-2 Neutralizing Antibody Geometric Mean Titres (PPI Subset)

| **Visit** | **Statistics** | **MVC-COV1901** | **Placebo** | **Ratio (MVC/Placebo)** | ***P*-value [1]** |
| --- | --- | --- | --- | --- | --- |
| **All Participants of PPI Subset, N** | | **903** | **150** |  |  |
| Day 1 (Vaccination 1) | n | 903 | 150 |  |  |
|  | GMT | 4.06 | 4.02 | 1.01 | 0.2406 |
|  | 95% CI of GMT | (4.02, 4.09) | (3.98, 4.07) | (0.99, 1.02) |  |
| Day 57 | n | 903 | 150 |  |  |
|  | GMT | 662.31 | 4.00 | 165.58 | <0.0001 |
|  | 95% CI of GMT | (628.66, 697.75) | (4.00, 4.00) | (157.16, 174.44) |  |
|  | n | 903 | 150 |  |  |
|  | GMT ratio | 163.22 | 0.99 | 164.15 | <0.0001 |
|  | 95% CI of GMT ratio | (155.01, 171.87) | (0.98, 1.01) | (155.71, 173.05) |  |
| **Subgroup: 20 to 64 years of age, N** | | **682** | **113** |  |  |
| Day 1 (Vaccination 1) | n | 682 | 113 |  |  |
|  | GMT | 4.06 | 4.03 | 1.01 | 0.4119 |
|  | 95% CI of GMT | (4.02, 4.11) | (3.97, 4.09) | (0.99, 1.03) |  |
| Day 57 | n | 682 | 113 |  |  |
|  | GMT | 732.89 | 4.00 | 183.22 | <0.0001 |
|  | 95% CI of GMT | (692.41, 775.74) | (4.00, 4.00) | (173.10, 193.94) |  |
|  | n | 682 | 113 |  |  |
|  | GMT ratio | 180.45 | 0.99 | 181.82 | <0.0001 |
|  | 95% CI of GMT ratio | (170.59, 190.89) | (0.98, 1.01) | (171.55, 192.70) |  |
| **Subgroup: ≥ 65 years of age, N** | | **221** | **37** |  |  |
| Day 1 (Vaccination 1) | n | 221 | 37 |  |  |
|  | GMT | 4.05 | 4.00 | 1.01 | 0.1730 |
|  | 95% CI of GMT | (3.98, 4.11) | (4.00, 4.00) | (0.99, 1.03) |  |
| Day 57 | n | 221 | 37 |  |  |
|  | GMT | 484.54 | 4.00 | 121.14 | <0.0001 |
|  | 95% CI of GMT | (433.16, 542.01) | (4.00, 4.00) | (108.29, 135.50) |  |
|  | n | 221 | 37 |  |  |
|  | GMT ratio | 119.75 | 1.00 | 119.75 | <0.0001 |
|  | 95% CI of GMT ratio | (107.16, 133.82) | (1.00, 1.00) | (107.16, 133.82) |  |

Abbreviations: N = number of participants in the population; n = number of participants with available data; GMT = geometric mean titre; CI = confidence interval

Note: Blood samples on Day 1 for immunogenicity test were collected before the administration of study intervention. GMT ratio was compared to Day 1 (prior to first dose).

[1] *P*-value based on two-sample t test or Wilcoxon rank sum test.

Table S6. Seroconversion Rate Based on the Wild Type SARS-CoV-2 Neutralizing Antibody Titres at Day 57 (PPI Subset)

| **Visit** | **Statistics** | **MVC-COV1901** | **Placebo** | **Treatment Difference % [1] (MVC-COV1901 minus Placebo)** | ***P*-value [2]** |
| --- | --- | --- | --- | --- | --- |
| **All Participants of PPI Subset, N** | | **903** | **150** |  |  |
| Day 57 | n | 903 | 150 |  |  |
|  | Seroconversion, n (%) | 901 (99.8) | 0 | 99.8 | <0.0001 |
|  | 95% CI | (99.20, 99.97) | (0.00, 2.43) | (97.51, 99.98) |  |
| **Subgroup: 20 to 64 years of age, N** | | **682** | **113** |  |  |
| Day 57 | n | 682 | 113 |  |  |
|  | Seroconversion, n (%) | 681 (99.9) | 0 | 99.9 | <0.0001 |
|  | 95% CI | (99.19, 100.00) | (0.00, 3.21) | (96.79, 100.00) |  |
| **Subgroup: ≥ 65 years of age, N** | | **221** | **37** |  |  |
| Day 57 | n | 221 | 37 |  |  |
|  | Seroconversion, n (%) | 220 (99.5) | 0 | 99.5 | <0.0001 |
|  | 95% CI | (97.50, 99.99) | (0.00, 9.49) | (90.46, 99.99) |  |

Abbreviations: N = number of participants in the population; n = number of participants in the specific category; %=percentage of participants with available data (n) as the denominator; CI = confidence interval.

Note: Seroconversion was defined as at least 4-fold increase of post-study intervention antibody titres from the baseline titre or from half of the lower limit of detection if undetectable at baseline. Blood samples on Day 1 for immunogenicity test were collected before the administration of study intervention.

[1] Treatment Difference was presented with the asymptotic 95% CI. In the case of small cell count (expected count less than 5), exact 95% CI was applied alternatively.

[2] *P*-value: Pearson's Chi-square test. In the case of small cell count (expected count less than 5), Fisher's exact test was applied alternatively.

Table S7. Anti-S IgG Geometric Mean Titres (PPI Subset)

| **Visit** | **Statistics** | **MVC-COV1901** | **Placebo** | **Ratio (MVC/Placebo)** | ***P*-value [1]** |
| --- | --- | --- | --- | --- | --- |
| **All Participants of PPI Subset, N** | | **903** | **150** |  |  |
| Day 1 (Vaccination 1) | n | 903 | 150 |  |  |
|  | GMT | 52.71 | 54.16 | 0.97 | 0.3870 |
|  | 95% CI of GMT | (51.65, 53.79) | (51.09, 57.40) | (0.92, 1.04) |  |
| Day 29 (Vaccination 2) | n | 901 | 147 |  |  |
|  | GMT | 430.48 | 55.68 | 7.73 | <0.0001 |
|  | 95% CI of GMT | (398.68, 464.83) | (51.71, 59.95) | (6.95, 8.60) |  |
|  | n | 901 | 147 |  |  |
|  | GMT ratio | 8.17 | 1.03 | 7.96 | <0.0001 |
|  | 95% CI of GMT ratio | (7.56, 8.82) | (0.96, 1.10) | (7.18, 8.82) |  |
| Day 43 | n | 898 | 149 |  |  |
|  | GMT | 8262.17 | 57.64 | 143.34 | <0.0001 |
|  | 95% CI of GMT | (7801.94, 8749.54) | (52.10, 63.76) | (127.65, 160.96) |  |
|  | n | 898 | 149 |  |  |
|  | GMT ratio | 156.70 | 1.06 | 147.31 | <0.0001 |
|  | 95% CI of GMT ratio | (147.52, 166.46) | (0.96, 1.17) | (131.31, 165.26) |  |
| Day 57 | n | 903 | 150 |  |  |
|  | GMT | 5745.35 | 54.43 | 105.56 | <0.0001 |
|  | 95% CI of GMT | (5464.49, 6040.64) | (51.66, 57.34) | (98.21, 113.46) |  |
|  | n | 903 | 150 |  |  |
|  | GMT ratio | 109.00 | 1.01 | 108.46 | <0.0001 |
|  | 95% CI of GMT ratio | (103.32, 115.00) | (0.96, 1.05) | (101.07, 116.38) |  |
| **Subgroup: 20 to 64 years of age, N** | | **682** | **113** |  |  |
| Day 1 (Vaccination 1) | n | 682 | 113 |  |  |
|  | GMT | 53.16 | 53.97 | 0.98 | 0.6524 |
|  | 95% CI of GMT | (51.87, 54.49) | (50.59, 57.58) | (0.92, 1.05) |  |
| Day 29 (Vaccination 2) | n | 682 | 112 |  |  |
|  | GMT | 524.24 | 55.98 | 9.36 | <0.0001 |
|  | 95% CI of GMT | (482.66, 569.41) | (51.10, 61.33) | (8.28, 10.59) |  |
|  | n | 682 | 112 |  |  |
|  | GMT ratio | 9.86 | 1.04 | 9.51 | <0.0001 |
|  | 95% CI of GMT ratio | (9.06, 10.73) | (0.95, 1.13) | (8.42, 10.75) |  |
| Day 43 | n | 679 | 113 |  |  |
|  | GMT | 9637.58 | 59.39 | 162.28 | <0.0001 |
|  | 95% CI of GMT | (9070.69, 10239.89) | (52.16, 67.61) | (138.63, 189.97) |  |
|  | n | 679 | 113 |  |  |
|  | GMT ratio | 181.24 | 1.10 | 164.71 | <0.0001 |
|  | 95% CI of GMT ratio | (169.83, 193.42) | (0.97, 1.25) | (142.79, 189.99) |  |
| Day 57 | n | 682 | 113 |  |  |
|  | GMT | 6520.95 | 54.79 | 119.02 | <0.0001 |
|  | 95% CI of GMT | (6188.26, 6871.32) | (51.47, 58.32) | (109.74, 129.10) |  |
|  | n | 682 | 113 |  |  |
|  | GMT ratio | 122.66 | 1.02 | 120.84 | <0.0001 |
|  | 95% CI of GMT ratio | (115.85, 129.88) | (0.96, 1.07) | (111.65, 130.79) |  |
| **Subgroup: ≥ 65 years of age, N** | | **221** | **37** |  |  |
| Day 1 (Vaccination 1) | n | 221 | 37 |  |  |
|  | GMT | 51.34 | 54.72 | 0.94 | 0.3584 |
|  | 95% CI of GMT | (49.67, 53.06) | (47.81, 62.64) | (0.82, 1.08) |  |
| Day 29 (Vaccination 2) | n | 219 | 35 |  |  |
|  | GMT | 233.05 | 54.70 | 4.26 | <0.0001 |
|  | 95% CI of GMT | (198.91, 273.05) | (48.89, 61.20) | (3.51, 5.16) |  |
|  | n | 219 | 35 |  |  |
|  | GMT ratio | 4.54 | 0.99 | 4.56 | <0.0001 |
|  | 95% CI of GMT ratio | (3.88, 5.31) | (0.94, 1.05) | (3.86, 5.40) |  |
| Day 43 | n | 219 | 36 |  |  |
|  | GMT | 5125.71 | 52.48 | 97.68 | <0.0001 |
|  | 95% CI of GMT | (4538.27, 5789.19) | (47.57, 57.89) | (83.67, 114.03) |  |
|  | n | 219 | 36 |  |  |
|  | GMT ratio | 99.81 | 0.96 | 104.35 | <0.0001 |
|  | 95% CI of GMT ratio | (87.87, 113.38) | (0.89, 1.02) | (90.42, 120.42) |  |
| Day 57 | n | 221 | 37 |  |  |
|  | GMT | 3887.09 | 53.35 | 72.86 | <0.0001 |
|  | 95% CI of GMT | (3476.08, 4346.71) | (48.46, 58.72) | (62.97, 84.31) |  |
|  | n | 221 | 37 |  |  |
|  | GMT ratio | 75.71 | 0.97 | 77.67 | <0.0001 |
|  | 95% CI of GMT ratio | (67.28, 85.20) | (0.90, 1.06) | (67.24, 89.71) |  |

Abbreviations: N = number of participants in the population; n = number of participants with available data; GMT = geometric mean titre; CI = confidence interval

Note: Blood samples or immunogenicity test were collected before the administration of study intervention. GMT ratio was compared to Day 1 (prior to first dose).

[1] *P*-value based on two-sample t test or Wilcoxon rank sum test.

Table S8. Seroconversion Rate Based on the Anti-S IgG Titres at Day 57 (PPI Subset)

| **Visit** | **Statistics** | **MVC-COV1901** | **Placebo** | **Treatment Difference % [1] (MVC-COV1901 minus Placebo)** | ***P*-value [2]** |
| --- | --- | --- | --- | --- | --- |
| **All Participants of PPI Subset, N** | | **903** | **150** |  |  |
| Day 29 (Vaccination 2) | n | 901 | 147 |  |  |
|  | Seroconversion, n (%) | 670 (74.4) | 2 (1.4) | 73.0 | <0.0001 |
|  | 95% CI | (71.38, 77.18) | (0.17, 4.83) | (68.83, 76.22) |  |
| Day 43 | n | 898 | 149 |  |  |
|  | Seroconversion, n (%) | 895 (99.7) | 2 (1.3) | 98.3 | <0.0001 |
|  | 95% CI | (99.03, 99.93) | (0.16, 4.76) | (95.11, 99.58) |  |
| Day 57 | n | 903 | 150 |  |  |
|  | Seroconversion, n (%) | 899 (99.6) | 0 | 99.6 | <0.0001 |
|  | 95% CI | (98.87, 99.88) | (0.00, 2.43) | (97.37, 99.89) |  |
| **Subgroup: 20 to 64 years of age, N** | | **682** | **113** |  |  |
| Day 29 (Vaccination 2) | n | 682 | 112 |  |  |
|  | Seroconversion, n (%) | 552 (80.9) | 2 (1.8) | 79.2 | <0.0001 |
|  | 95% CI | (77.79, 83.82) | (0.22, 6.30) | (74.09, 82.70) |  |
| Day 43 | n | 679 | 113 |  |  |
|  | Seroconversion, n (%) | 677 (99.7) | 2 (1.8) | 97.9 | <0.0001 |
|  | 95% CI | (98.94, 99.96) | (0.22, 6.25) | (93.68, 99.48) |  |
| Day 57 | n | 682 | 113 |  |  |
|  | Seroconversion, n (%) | 679 (99.6) | 0 | 99.6 | <0.0001 |
|  | 95% CI | (98.72, 99.91) | (0.00, 3.21) | (96.63, 99.91) |  |
| **Subgroup: ≥ 65 years of age, N** | | **221** | **37** |  |  |
| Day 29 (Vaccination 2) | n | 219 | 37 |  |  |
|  | Seroconversion, n (%) | 118 (53.9) | 0 | 53.9 | <0.0001 |
|  | 95% CI | (47.04, 60.62) | (0.00, 10.00) | (42.11, 60.82) |  |
| Day 43 | n | 219 | 36 |  |  |
|  | Seroconversion, n (%) | 218 (99.5) | 0 | 99.5 | <0.0001 |
|  | 95% CI | (97.48, 99.99) | (0.00, 974) | (90.21, 99.99) |  |
| Day 57 | n | 221 | 37 |  |  |
|  | Seroconversion, n (%) | 220 (99.5) | 0 | 99.5 | <0.0001 |
|  | 95% CI | (97.50, 99.99) | (0.00, 9.49) | (90.46, 99.99) |  |

Abbreviations: N = number of participants in the population; n = number of participants in the specific category; %=percentage of participants with available data (n) as the denominator; CI = confidence interval.

Note: Seroconversion was defined as at least 4-fold increase of post-study intervention antibody titres from the baseline titre or from half of the lower limit of detection if undetectable at baseline. Blood samples for immunogenicity test were collected before the administration of study intervention.

[1] Treatment Difference was presented with the asymptotic 95% CI. In the case of small cell count (expected count less than 5), exact 95% CI was applied alternatively.

[2] *P*-value: Pearson's Chi-square test. In the case of small cell count (expected count less than 5), Fisher's exact test was applied alternatively.

### Supplementary Figures

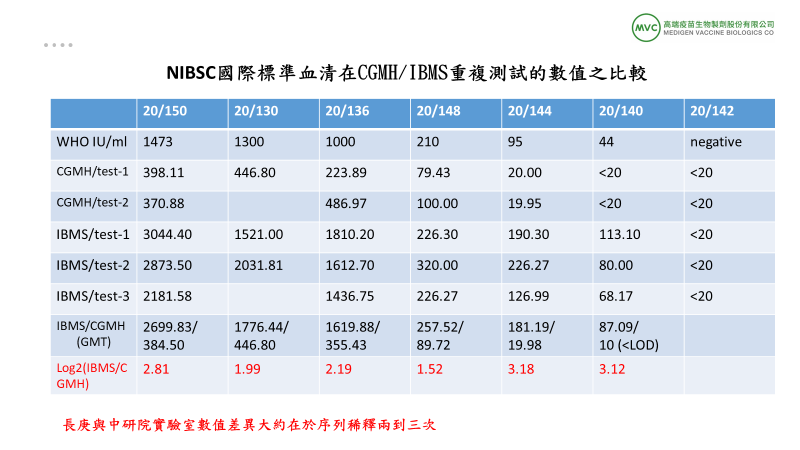

#### Fig S1. Reporting Neutralizing Antibody Titres using the WHO International Unit (IU/mL)

To establish the reporting of neutralizing antibody titres in our central labs, CGMH and IBMS, we purchased the WHO standardized sera, the research reagent 20/130, WHO International Standard 20/136 and the WHO Reference Panel (20/150, 20/148, 20/144, 20/140, and 20/142) from NIBSC. Each standardized serum was marked with the arbitrarily assigned unitage, IU/mL. These sera were tested in the neutralizing antibody assays in the two labs along with other samples to obtain the NT50 values for equation establishment.

The neutralizing antibody titres of the respective sera were obtained through repeated tests. We repeated the test twice at CGMH (phase 1 lab) and three times at IBMS (phse 2 lab). The raw data were listed as Fig S1. Taking the difference of log2 of the NT50 value, it is estimated that the two labs had 2 to 3 difference in interms of number of serial dilution.
